# Healthcare use and cost trajectories during the last year of life: A national population administrative secondary care data linkage study

**DOI:** 10.1101/2020.09.29.20203794

**Authors:** Katharina Diernberger, Xhyljeta Luta, Joanna Bowden, Marie Fallon, Joanne Droney, Elizabeth Lemmon, Ewan Gray, Joachim Marti, Peter S Hall

## Abstract

**Background:** People who are nearing the end of life are high users of healthcare. The cost to providers is high and the value of care is uncertain.

**Objectives:** To describe the pattern, trajectory and drivers of secondary care use and cost by people in Scotland in their last year of life.

**Methods:** Retrospective whole-population secondary care administrative data linkage study of Scottish decedents of 60 years and over between 2012 and 2017 (N=274,048).

**Results:** Secondary care use was high in the last year of life with a sharp rise in inpatient admissions in the last three months. The mean cost was £10,000. Cause of death was associated with differing patterns of healthcare use: dying of cancer was preceded by the greatest number of hospital admissions and dementia the least. Greater age was associated with lower admission rates and cost. There was higher resource use in the urban areas. No difference was observed by deprivation.

**Conclusions:** Hospitalisation near the end of life was least frequent for older people and those living rurally, although length of stay for both groups, when they were admitted, was longer. Research is required to understand if variation in hospitalisation is due to variation in the quantity or quality of end of life care available, varying community support, patient preferences or an inevitable consequence of disease-specific needs.

## Background

Improving the availability and quality of palliative and end of life care is a global priority set out by the WHO in their resolution on palliative care.^1^ In 2015 The Scottish Government published The Strategic Framework for Action for Palliative and End of Life Care, with a vision that ‘*By 2021, everyone in Scotland who needs palliative care will have access to it*.’ ^2^ Whilst ambitious, the vision was said to be achievable through commitments that included commissioning guidance for health and social care partnerships and research to understand current unmet needs and unwarranted variation in access to the right care and outcomes.

A systematic review of Scotland-based palliative care research published in 2018 revealed a lack of health economic research. ^3^ This was a timely observation with growing interest in demonstrating the value of healthcare, from the perspectives of people receiving care, and on the part of service commissioners and providers. Realistic Medicine, a landmark report from Scotland’s Chief Medical Officer published in 2015, provided clear expectations of a future healthcare system that offered true value and minimised waste; with ‘waste’ described from the healthcare recipient’s perspective, as interventions that do not add value to their care. ^4,5^

People who are nearing the end of life are high users of secondary care services. Around 50% of people in Scotland currently die in hospital.^6,7,8^ Hospitalisation may be odds with the expressed preferences of people living with advanced illness^9^. It may be recommended for some people with complex clinical needs, but may also represent a culture and associated practices of so-called ‘over-medicalisation’; whereby hospital-based care and interventions do not offer meaningful benefit to individuals and may even cause harm.^10,11^

The Community Health Index (CHI) number is a unique identifier allocated to every Scottish resident offers a unique opportunity for robust population-based data linkage. ^12^ It has allowed Scotland to be at the forefront of population research in this area with recent studies showing that both time-to-death age at death and socioeconomic status are significant predictors of cost in the last three years of life. ^13, 14^

Understanding the type, intensity of care and variation that people nearing the end of life receive is an important before recommendations can be made to improve access to appropriate palliative care.^15^

There were three key objectives:

1. To describe secondary healthcare use, trajectory and associated costs over the last year of life for the Scottish population.
2. To describe patterns of healthcare use for disease-specific subpopulations.
3. To investigate associations between demographic characteristics, including age, and secondary healthcare access in the last year of life, in order to highlight possible unwarranted variation.

## Methods

A retrospective population-level data linkage study was undertaken, including all decedents in Scotland in 2012 – 2017, who were over 60 years of age on their date of death. Secondary healthcare use was examined over the last 12 months of life. Deaths under the age of 60 were not included, in order to maintain sufficient underlying disease prevalence for meaningful study.

### Data sources

Data were obtained from Public Health Scotland via the Scottish Research Data Safe Haven. Linkage was established between the Scottish Morbidity Record (SMR) outpatient, inpatient and day case and the National Records of Scotland (NRS) record of deaths using CHI number as the primary key for linkage.^9^ SMR01 includes episode-based patient records that relate to all acute inpatient and day cases. To reduce measurement error, the SMR01 data were checked for data entry anomalies such as duplicates, overlapping and nested episodes (See Supplementary table 1). SMR00 relates to all outpatients (new and follow-up) in specialties other than Accident & Emergency (A&E) and Genito-Urinary Medicine. In addition, we relied on NRS-deaths data and the SIMD.

### Inclusion and exclusion criteria

Detailed eligibility criteria are reported in Figure S1 (supplementary material). Major inclusion criteria were:

- Death registered between January 1^st^ 2012 and December 31^st^ 2017
- Age at death ≥60 years
- Healthcare data available for a minimum of 12 months prior to death

Figure S1 describes the data sources and the selection of the study population. The NRS death dataset of participants meeting the eligibility criteria was merged with the outpatient dataset SMR00 and with the inpatient and day case data SMR01. Inpatient and outpatient resource use was excluded if the patient identifier (PID) was missing or if the resource use occurred outside the study period.

### Patient characteristics

Patient characteristics included gender, age and primary cause of death (one of five ICD-10 categories Cancer, Circulatory, Respiratory, Dementia and other). Comorbidity was estimated using the Charlson Comorbidity Index (CCI). ^16^ The CCI was based on secondary care coding from the last year of life, with the limitation that only patients who accessed secondary care during their last year of life had a CCI score. An urban-rural indicator was included, developed by the Rural and Environment Science and Analytical Services Division and the Scottish index of Multiple Deprivation (SIMD). ^17^

### Outcome measures

#### Inpatient and Day Care

Hospital inpatient care in the last year of life was captured as: number of hospital admissions, mean number of bed days per stay and total number of bed days over the 12 month period.

To estimate the cost of inpatient care, the Scottish health service costs (Scottish cost book) was used, mainly R040 (Specialty group costs-Inpatients in all specialties excluding long stays) and R040LS (Specialty group costs-Inpatients in all specialties long stays). Critical care stays are included within mean costs. Day cases were costed using R042 (Specialty group costs-day cases).^18^

#### Outpatient care

Outpatient data included the number and nature of outpatient appointments per patient in the last year of life. Costs for outpatient appointments were derived from the Scottish health service costs documents R044 (Specialty group costs-consultant outpatients), R045 (Specialty group costs-Nurse led clinics) and R046 (Specialty group costs – Allied Health Professionals). The costs are based on national average unit costs for each service code.

### Statistical Analysis

Descriptive statistics were used to characterise the study population. Means and standard deviation (SD) were calculated for services and costs. Generalised linear models (GLM) were used to estimate costs, adjusting for age, gender, primary cause of death, SIMD, an urban-rural indicator and comorbidity.^19,20^ We also assessed potential interactions between age and gender as well as age and cause of death. Analysis used Stata version 16 (StataCorp, College Station, TX, USA).

### Ethics

Approval was granted by the Scottish Public Benefit and Privacy panel (Ref: 1617-0100) for analysis within the Scottish National Research Data Safe Haven.

## Results

### Patient characteristics

A total of 339,963 people died in Scotland between January 1^st^ 2012 and December 31^st^ 2017, of whom 274,048 met the eligibility criteria.

Sixty percent of the decedent population were under 80 years old at death and 54% were female (Table 1). The most common causes of death were circulatory diseases (29.2%) and cancer (27.5%). Around two thirds of the study population lived in urban areas, around 20% in accessible small towns or accessible rural areas and 10% in small remote towns and remote areas.

**Table 1:**
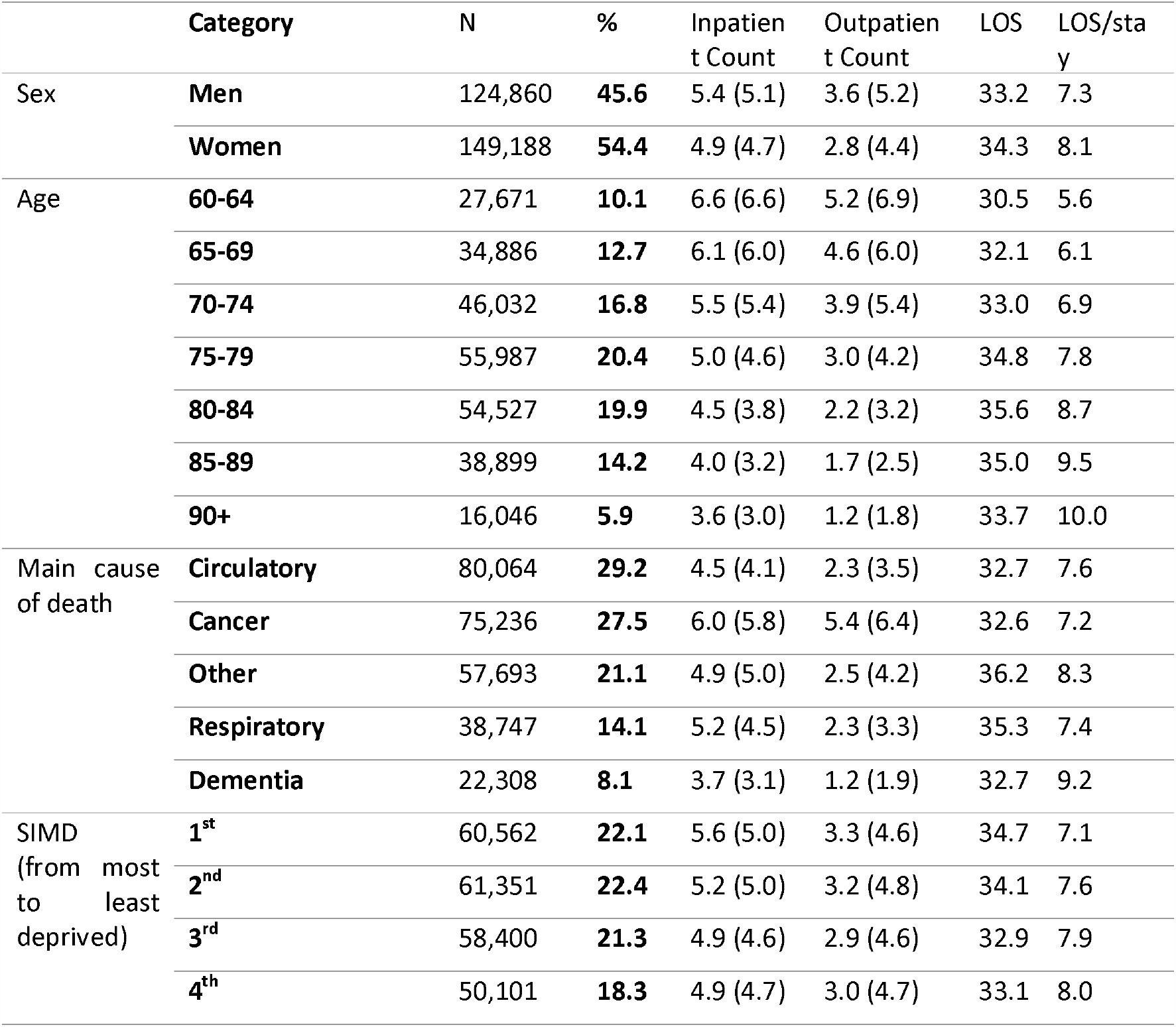
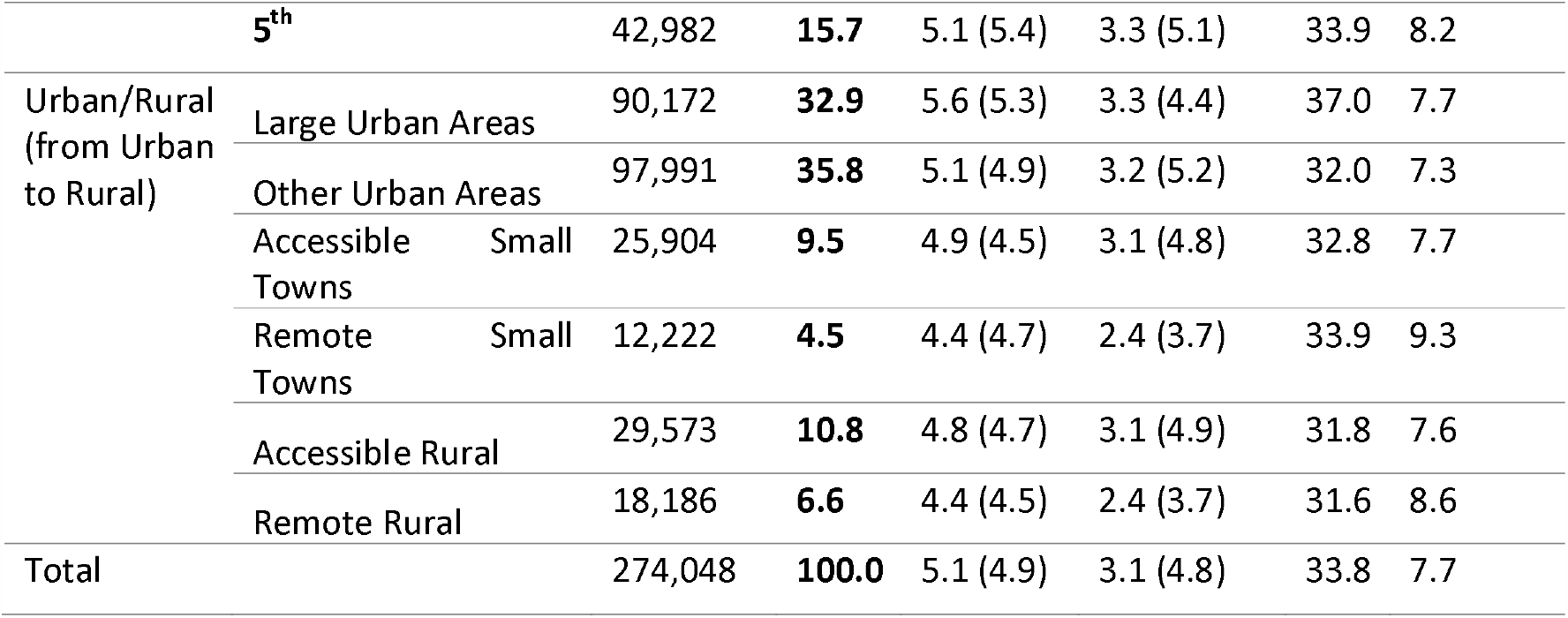
Study population and resource use [outpatient and inpatient count and standard deviation, overall length of stay (LOS) within a hospital and average in-hospital time per inpatient stay] in the last year of life

### Inpatient, outpatient and day case use and costs

The mean number of hospital inpatient admissions during the last year of life was 5.1 (SD: 4.9) and hospital outpatient appointments 3.1 (SD: 4.8) (Table 2). The mean total number of hospital bed days in the last year life was 33.8, with a mean length of stay per admission of 7.7 days. Examining the unadjusted differences, males had a higher number of inpatient and outpatient appointments than females but spent fewer total days in hospital due to a shorter average length of stay. Around three-quarters of study participants were hospitalized at least once in their last year, and 29% in their last month of life. Nearly 80% of the study population had one or more outpatient appointment in their last year, with one-third having an outpatient appointment during their last month of life. The number of day case appointments was comparably small with 6.3% of the population having one or more in their last year and just under 0.5% having a day case appointment in the last month of life.

**Table 2:** Health Care utilisation in the last year and in the last month of life

| Healthcare Utilisation | N = 274048 | Last 12 month | Last month |
| --- | --- | --- | --- |
| **Inpatient care** | Hospitalizations, No.% | 200510 (73.16%) | 80287 (29.296%) |
|  | Hospital admissions, mean (SD) | 5.1 (4.9) | 0.856 (1.399) |
|  | Hospital duration(days), mean (SD) | 33.8 (37.8) | 5.14 (6.48) |
| **Day cases** | Day cases, No.% | 17540 (6.3%) | 1363 (0.497%) |
| **Outpatient care** | Outpatient visits, No. (%) | 217772 (79.465%) | 92454 (33.736%) |
|  | Outpatient visits, mean (SD) | 3.1 (4.8) | 0.406 (0.7007) |

The mean cost of secondary care was £ 10,134 (CI [9,921, 10,337]) per person in the last year of life (Supplementary table S3). Proximity to death had the biggest influence on adjusted monthly costs (Supplementary table S4). The main contributor to costs over the final 12 months of life was inpatient hospital stays; peaking during the last three months when admissions were most common.

### Healthcare use by primary cause of death

There were significant differences in patterns of healthcare use by decedents’ cause of death. (Figure 1). Inpatient hospitalisation rates accelerated over the last year of life for all causes of death, and this was most pronounced in circulatory or respiratory disease. Patients who died from cancer accessed more day care over the last year of life. Frequency of outpatient care remained relatively constant over the last year of life for most groups, except for those who died of cancer were higher users of all three domains of secondary healthcare.

**Figure 1:**
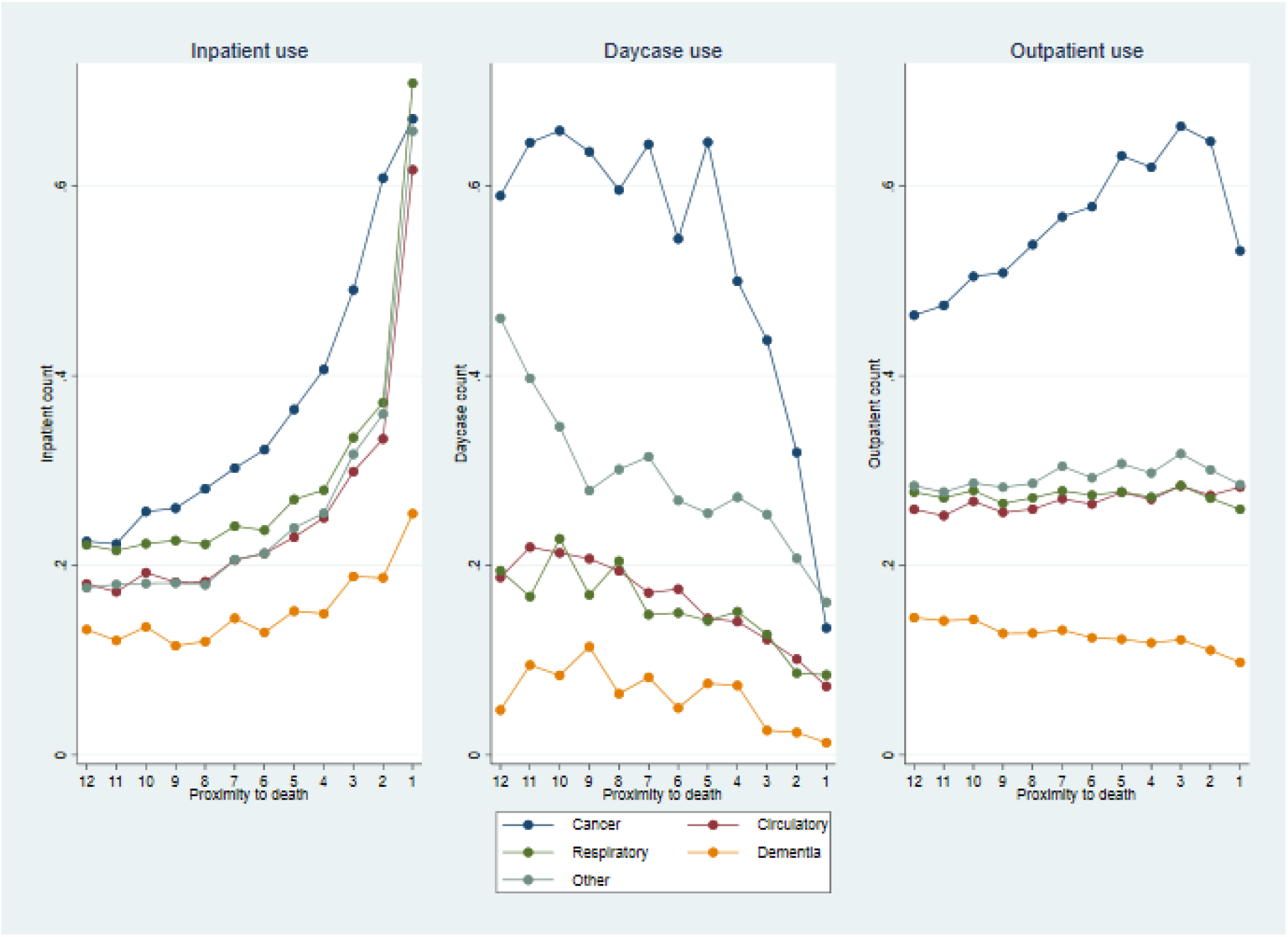
Resource utilisation (secondary care) in the last year of life for inpatient, day case and outpatient use split by main causes of death

218,357 decedents had an evaluable CCI score, with missing data reflecting those with no hospital records during the last year of life. Around one third of this sub-population had a CCI score of zero, 60% had a score between 1 and 5 and the remaining 5% had a score between 6 and 12 reflecting the highest disease burden (Supplementary table 2). Adding comorbidity as an explanatory variable into a GLM (Supplementary table 3) it can be observed that a higher CCI is associated with higher secondary healthcare costs, but with some variability.

### Healthcare use and costs by age and demographics

Inpatient hospitalisation increased in frequency in all age categories with proximity to death, with a steep rise in the last three months of life (Figure 2). The frequency of day case use varied considerably over the last year for all age groups, though younger patients accessed significantly more day case care. Outpatient use was largely constant across the last year for the population groups over 75 years of age, whilst the younger population had a slight increase until three months prior to death, followed by a sharp decrease.

**Figure 2:**
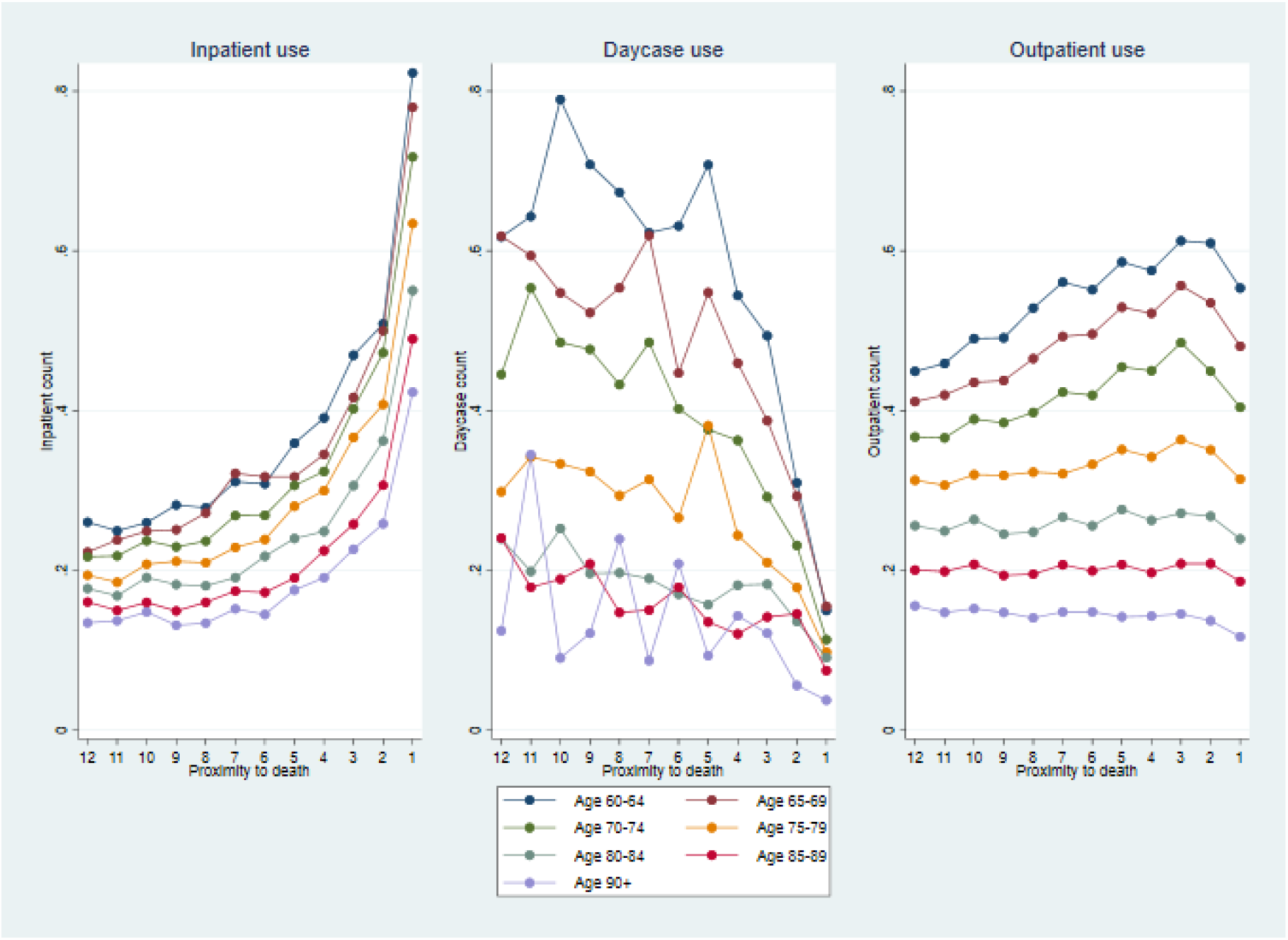
Resource utilisation in the last year of life for inpatient, day case and outpatient use split by age groups. (the y-axis displays the average number of inpatient, day case and outpatient appointments in the specified month)

One year adjusted and unadjusted costs decreased with increasing age (Table 3 and Figure 2). Unadjusted costs for the youngest group were £12,420.7, which was double the costs for those aged 90 and over. However, after adjusting for gender, primary cause of death, SIMD, RU and comorbidity, costs for the youngest and the oldest begin to converge.

**Table 3:** Generalised linear model - unadjusted costs with CIs on the left, adjusted generalised linear model on the right

| Age group | Unadjusted Costs (£) | CI (95%) | Adjusted Costs (£) | CI (95%) |
| --- | --- | --- | --- | --- |
| **60-64** | **12420.7\*\*\*** | [12262.5,12578.9] | **12411.4\*\*\*** | [12271,12551,9] |
| **65-69** | **11522.1\*\*\*** | [11391.9,11652.4] | **11894.1\*\*\*** | [11775.6, 12012.5] |
| **70-74** | **10695.5\*\*\*** | [10590.5,10800.4] | **11615.5\*\*\*** | [11515.5, 11715,4] |
| **75-79** | **9525.4\*\*\*** | [9440.4,9610.3] | **11331.6\*\*\*** | [11240.7, 11422,5] |
| **80-84** | **8406.5\*\*\*** | [8330.1,8482.9] | **10970.3\*\*\*** | [10876.9, 11063.7] |
| **85-89** | **7296.2\*\*\*** | [7216.6,7375.7] | **10499.6\*\*\*** | [10388.8, 10610.4] |
| **90+** | **6117.3\*\*\*** | [6009.0,6225.5] | **10019.3\*\*\*** | [9843.35, 10195.4] |
| **Observations** | 260636 |  |  |  |
| Adjusted for gender, main cause of death, SIMD, Urban-rural Indicator, Charlson index |  |  |  |  |
| 95% confidence intervals in brackets * $p < 0.05$, ** $p < 0.01$, *** $p < 0.001$ | | | | |

People living in large urban areas had highest use of all types of healthcare (Figure 3). Those in remote small towns and remote rural areas used fewest resources, with exceptionally low use of outpatient appointments. No clear trend was observed with deprivation presented in Supplementary table 2.

**Figure 3:**
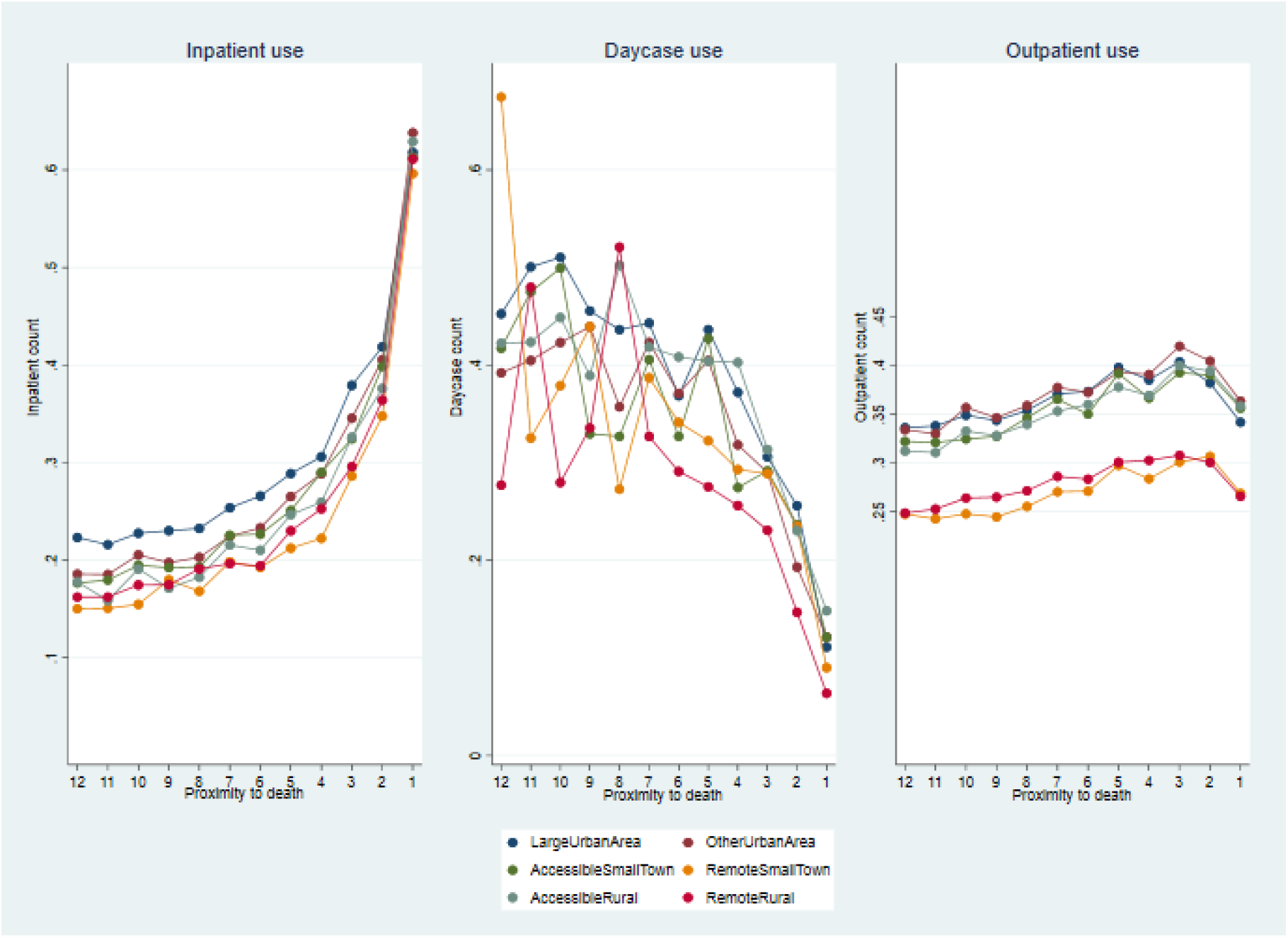
Resource utilisation in the last year of life for inpatient, day case and outpatient use split by rural-urban indicator

## Discussion

### Main findings

Inpatient hospitalisation was increasingly common over the last year of life and particularly when close to death. This was consistent across all causes of death, age and rurality groups. On average, people spent more than one of their last 12 months of life in hospital, typically over several admissions. Inpatient costs comprised the greatest proportion of the £10,000 average secondary care cost, at more than £8,500 per decedent.

The intensity and pattern of daycase care and outpatient appointments was more mixed, although the use of both fell sharply close to death with considerably smaller costs, at £400 and £650 respectively.

### Strengths and limitations of the study

The primary strength of our study is that our data covered the entire Scottish population of decedents and captured near-complete secondary care use during their last year of life. Therefore, our resource use estimates are less prone to bias due to non-random selection, as may occur in cohort studies.

The main limitation of our study was the scope of available data. Community palliative care services data was not available. We further acknowledge that secondary care represents only one dimension of healthcare and that primary care and social care are critical pieces of the care jigsaw. Inference can be made through join consideration with our parallel publication of English data that included primary care [EDITOR MAY CROSS-REFERENCE]

### What this study adds

Primary cause of death was clearly associated with differing patterns of healthcare use. The population who died of cancer were consistently the most frequent users of secondary healthcare, with those dying of dementia consistently the least following a pattern laid out by Murray et al.^21^ Older decedents used significantly less secondary healthcare during their last year of life, as did those living rurally.

The extent to which observed patterns of use reflect the needs or preferences of the different populations is unknown. Further research is needed to explore this and to investigate the likelihood of benefit of secondary care interventions close to death. This would allow quantification of the value of care.

Patterns of healthcare use are inevitably influenced by clinical service configuration. For instance, cancer care is predominantly secondary care outpatient-led, with individuals typically receiving treatment as day cases. Therefore, it is not surprising that outpatient and day case use was observed to be particularly high in this subpopulation. Services for people with dementia are more likely to be community or social care-based and it follows that this population access secondary care less than other groups.

The accessibility of healthcare is important, highlighted by our finding that rural populations access lower levels of secondary healthcare during their last year. We do not know whether rural individuals access more primary care or indeed whether their needs differ from those in more urban areas. Research is needed to explore the extent to which this, and other observed variations, are warranted.

### Conclusion

Improving the quality and appropriateness of care for people in the last phase of life is a national and international priority.^1,2,5^ We have described patterns of secondary healthcare use and associated costs for over a quarter of a million Scottish decedents; highlighting most significantly that inpatient hospitalisation accounts for a great proportion of costs, and is of uncertain value.

Detailed prospective quantitative and qualitative exploration around the value of admissions, day care and outpatient visits in the last year of life is needed. Apart from crucial insight into the patient experience and appropriateness of care, this could illuminate the realities of gaps in care and inequalities.

We require better insight into the value of the social care system and how community care can be a realistic alternative to hospital-based care. ^22^ Integrated health and social care in Scotland^2^is a new reality and provides opportunity for whole system learning.^2^

Ultimately, our goal must be to maximise value all round, with people nearing the end of life receiving high value care that is tailored to their needs, but simultaneously offers value to care commissioners and providers.

## Data Availability

Data are not available for sharing via application to the Scottish Public Benefits and Privacy Panel.

## Acknowledgements

The authors acknowledge the support of the Electronic Data Research and Innovation Service (eDRIS) team (Public Health Scotland) for their involvement in obtaining approvals, provisioning and linking data and the use of the secure analytical platform within the National Safe Haven. Further the authors would like to thank all members of the scientific advisory board namely: Julia Riley, Sandra Campbell, Catherine Urch, Bee Wee, Harry Quilter-Pinner, Ivor Williams and Gianluca Fontana for their valuable input.

## Authorship

JM and PH led the conception and design of the study. KD led the data acquisition, conducted data management and analysis supported by XL, EG, JM and PH. KD and EG led the data interpretation supported by XL, JB, EL, JM and PH. KD drafted and revised the article supported by JB and PH. All authors critically reviewed and edited the paper and approved the final version to be published.

## Patient consent

Not required

## Provenance and peer review

Not commissioned; externally peer reviewed.

